# Factors influencing the prescription of first-line treatment for type 2 diabetes mellitus: a systematic review

**DOI:** 10.1101/2025.01.22.25320805

**Authors:** Helena Moreira, Fernando Moreira, Ângelo Jesus, Matilde Monteiro-Soares, Paulo Santos

## Abstract

**Introduction:** Understanding and predicting prescription preferences for type 2 diabetes mellitus, a heterogeneous and complex condition that affects over 10% of the global adult population, can improve prescribing practices, guide policymakers in promoting evidence-based medicine, and help tailor first-line treatments to individual characteristics or specific subgroups, improving patient outcomes. This study aimed to identify factors influencing metformin prescription, the first-line therapy recommended in Western guidelines, and to assess factors leading to its avoidance and their alignment with evidence-based medicine. It also explores factors associated with initial combination therapy, a newer and controversial approach compared to stepwise therapy.

**Methods:** We performed a systematic search in PubMed, Scopus and Web of Science for observational analytical studies evaluating factors associated with metformin or combination therapy initiation. Quality assessment was done using the Joanna Briggs Institute critical appraisal checklists. (PROSPERO registry number CRD42023438313).

**Results:** Thirty studies were included, evaluating 105 variables, mostly (62%) assessed in one study. The 25 variables using combination therapy as outcome were mostly (72%) evaluated also in one study. Initial metformin prescription was strongly associated with the age of individuals with diabetes, glycated haemoglobin levels, body mass index, and renal complications, while combination therapy was mainly linked with glycated haemoglobin levels and comorbidities. Findings also highlighted a discrepancy between clinical practice and evidence-based recommendations. However, concerns were raised regarding both the internal and external validity of the included studies.

**Conclusion:** Our systematic review, that offers insights into real-world clinical practices, indicated that there is a misalignment between clinical practices and evidence-based recommendations supporting the need for interventions of this field.

## Introduction

More than 90% of people with diabetes have type 2 diabetes mellitus (T2DM), which is a chronic complex disease, and management requires a multifactorial approach to prevent or delay microvascular and macrovascular complications [1]. With a global prevalence of 10.5% among individuals aged 20 to 79 in 2021 [2], T2DM contributes to 11.3% of deaths worldwide [3]. It has led to a 315% increase in healthcare expenditures over 15 years (2007-2021) [2], significantly burdening healthcare systems and society.

Several classes of antidiabetic drugs (ADs) are currently available, each with distinct profiles of effectiveness and safety [4]. The most commonly used ADs belong to seven drug groups: biguanides, sulfonylureas (SU), thiazolidinediones (TZDs), dipeptidyl peptidase-4 inhibitors (DPP4i), sodium-glucose transporter-2 inhibitors (SGLT2i), glucagon-like peptidase receptor-1 agonists (GLP1-RA) and insulins [5]. Metformin (biguanide) is a unanimous recommendation as first-line in Western guidelines[4, 6, 7]. However, the American Diabetes Association (ADA) and the European Association for the Study of Diabetes (EASD) acknowledge that alternative first-line treatments may be appropriate. Specifically, SGLT2i and GLP1-RA are recommended for individuals with established cardiovascular disease, high cardiovascular risk, heart failure, or chronic kidney disease (CKD), irrespective of metformin use. Further, they also suggest initial combination therapy (i.e., starting with two or more drugs) when the glycated haemoglobin (HbA1c) is 1.5% above the target at diagnosis (i.e., >70 mmol/mol [>8.5%]) [4]. Therefore, with multiple therapeutic options available for treating this complex chronic disease, selecting the appropriate drug(s) can be a challenge for physicians.

Variation in healthcare resources and utilization raises essential questions about resource allocation quality, equity, and efficiency, with significant implications for healthcare and health policy [8]. Such variation is more likely to occur when multiple treatment options are available, leading to uncertainty about the most appropriate clinical choice, a phenomenon known as “professional uncertainty” [9]. Given that diabetes is a complex medical condition with diverse clinical manifestations and varying treatment responses, variations related to clinical conditions and individual preferences are expected. However, how much of this variation can be attributed to factors other than differences in individual health status or preferences?

A systematic review that approaches factors influencing first-line choice decisions in T2DM has yet to be found. Such a review would offer a more comprehensive perspective on potential factors and demonstrate the robustness of scientific evidence in this field. Further, identifying the factors can reveal possible clinical variations and lead to actions that promote more evidence-based medicine and increase equity in healthcare. Given that metformin is the recommended first-line treatment in guidelines and that combination therapy, which may include metformin, is a newer approach, this systematic review aims to summarize the key predictors for prescribing metformin or combination therapy as the first-line treatment for T2DM.

## Material and Methods

This systematic review strictly adhered to the Preferred Reporting Items for Systematic Reviews and Meta-Analyses (PRISMA) guidelines [10] and was duly registered in the PROSPERO database in July 2023 under CRD42023438313. The protocol registered in PROSPERO outlines a more comprehensive systematic review, part of which is presented here.

### Search strategy

A search was conducted in Medline (PubMed), Scopus and Web of Science on August 25, 2023, without language or time restrictions. With the clinical question (PECO-S) formulated, the search strategy was developed, with the Population (individuals with T2DM drug-naïve), Exposure (predictive factors), Outcome (starting metformin or combination therapy), and type of Study (observational analytical studies). The search strategy combined medical subject headings, free terms, and terms in the title/abstract, and it is available in S1, S2, and S3 Tables. Further, the references of relevant articles were manually searched for potentially eligible studies, and experts were also consulted. The grey literature was searched through ProQuest, Networked Digital Library of These and Dissertations, Eldis, and targeted websites like the World Health Organization (WHO), the United Nations, the International Diabetes Federation, the New York Academy of Medicine, and the National Institutes of Health.

### Inclusion and exclusion criteria

The inclusion and exclusion criteria were selected based on the PECO-S elements, and all details are shown in Table 1.

**Table 1:** Study inclusion and exclusion criteria.

| Category | Inclusion criteria |
| --- | --- |
| Study population | At least 80% adults ($\geq 18$ years old) with T2DM and naïve to antidiabetic medications or subgroup analysis provided. |
| Expose/Exhibit | All factors that can influence prescribing, such as physician-related factors, patient-related factors, healthcare system-related factors, influence of pharmaceutical companies and cost. |
| Outcomes | The choice of first-line oral antidiabetic therapy: metformin (a biguanide) or combination therapy (i.e., starting with two or more drugs simultaneously). |
| Publication type | Observational analytical studies. |
| Category | Exclusion criteria |
| Study population | Studies on pregnant or breastfeeding women. |

### Study selection

EndNote 20® was used to manage references and identify duplicates. After that, the Rayyan QCRI [11] tool was employed to allow two researchers (MH and MF or JA) to blindly and independently select the references for this systematic review. Titles and abstracts were screened in the first stage, with any disagreements between researchers leading to studies being transferred to the second stage. In the second stage, the full text was analysed. Disagreements were resolved by consensus; a third researcher was consulted if a consensus was not reached. This procedure was done for study selection, data extraction and risk of bias analysis. The agreement proportion between the two reviewers was calculated for each stage.

### Data extraction

Data were extracted by MH and confirmed by MF or JA following previously determined variables: (1) study identification (title, author(s), publication year, and country), (2) methods (sources and methods of participants selection, inclusion and exclusion criteria, sample size and its characteristics (gender and mean age), period(s) of data collection, follow-up, setting, statistical analysis methods, and potential bias), (3) outcomes analysed and their prevalence, list of variables (factors) and degree of statistical significance associated with the outcomes.

### Risk of bias and data analysis

Two independent reviewers (MH and MF or JA) assessed the quality of studies using the Joanna Briggs Institute (JBI) quality assessment checklist for cross-sectional and cohort studies[12].

Due to the expected and observed high heterogeneity of studies, a meta-analysis was not feasible. Instead, a narrative synthesis occurred to map all predictive factors influencing metformin or combination therapy as a first-line choice. The criterion for a statistically significant association between exposure and outcomes was p-value <0.05, or a 95% confidence interval that did not include 1 in studies where the association was reported through risk measures (relative risk, odds ratio or hazard ratio).

## Results

### Study selection

Fig 1 presents the selection process flowchart providing the main reasons for exclusion. Initially, 1,645 studies were identified, and four additional studies were found through reference reviews of the included studies. In the end, after two stages of screening studies, 30 studies were included. The proportion of agreement between the independent assessors from the first and second stages of screening studies was 51.4% and 69.5%, respectively.

**Fig 1:**
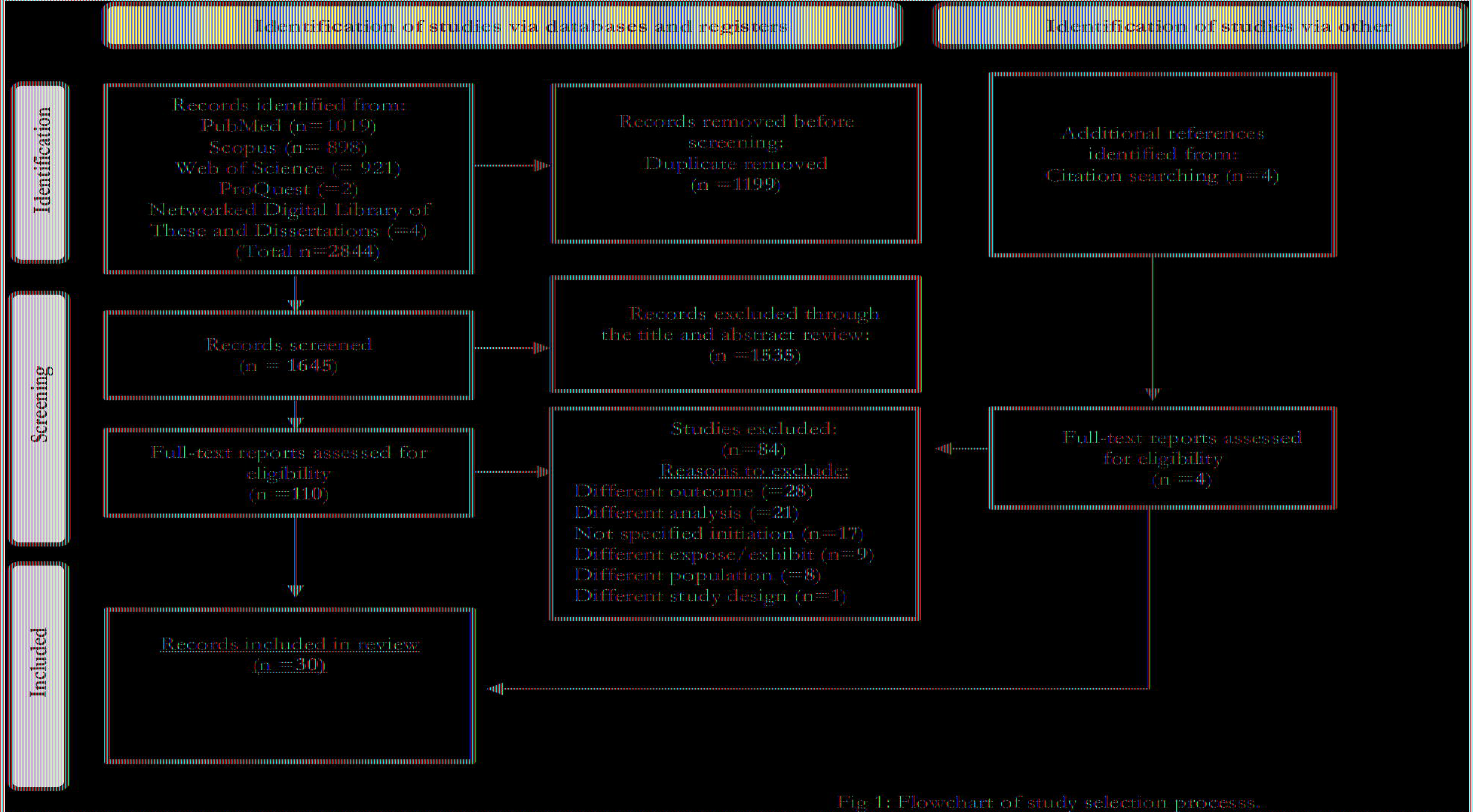
Flowchart of study selection

### Study characteristics

Tables 2 and 3 presents the characteristics of the included studies. Considering the study design, 9 (30%) [13–21] were retrospective cohort studies and 21 (70%) [22–42] were cross-sectional studies. Most studies were from Europe (n=12 (40%)), North America (n = 11 (36.7%)), with only 6 (20%) being from Asia and 1 (3.3%) from Australia. The number of participants varied widely, ranging from 415 to 1,136,723, with a median of 27,138.

**Table 2:**
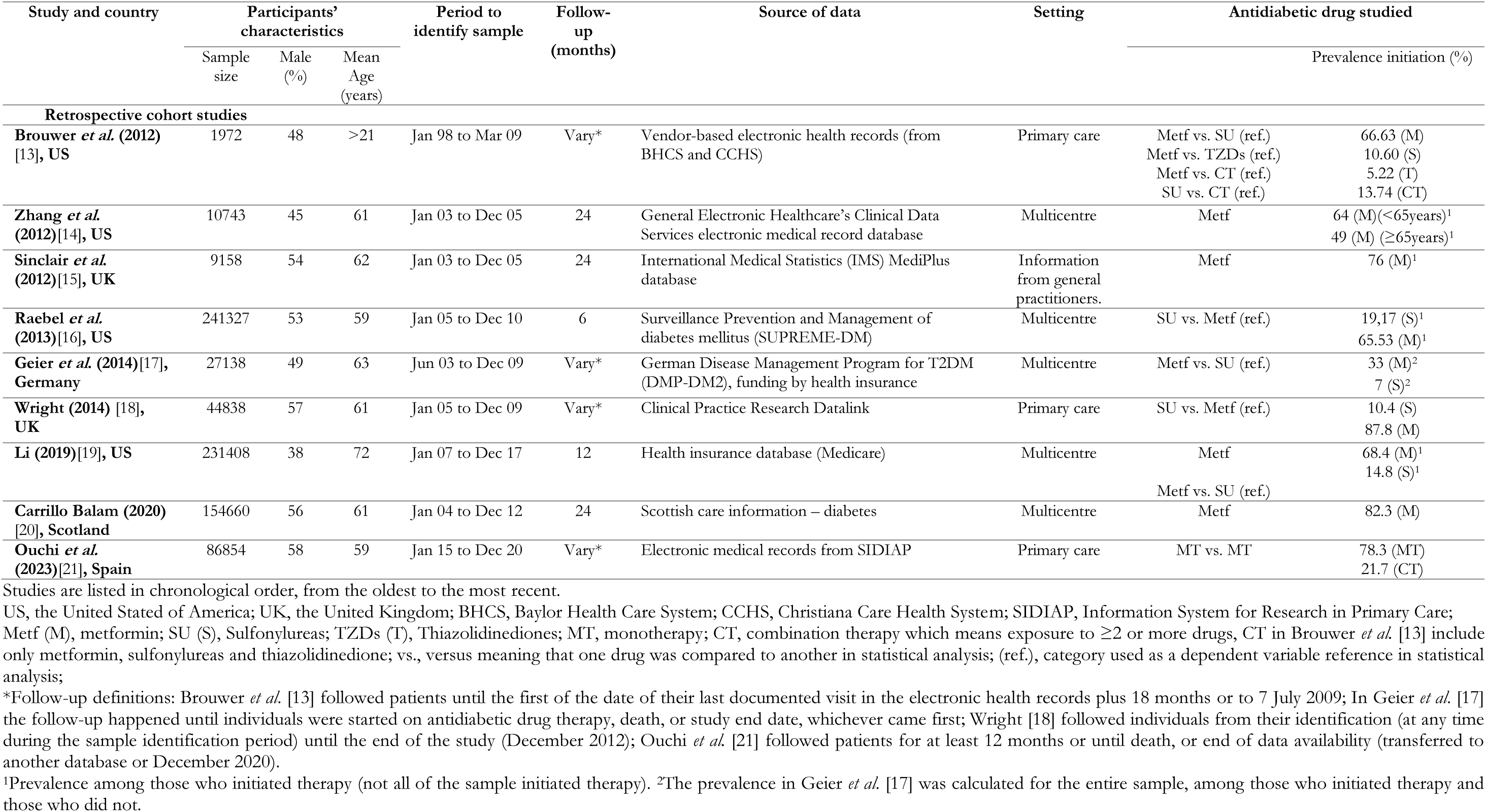
Characteristics of the retrospective cohort studies which were eligible for inclusion.

**Table 3:**
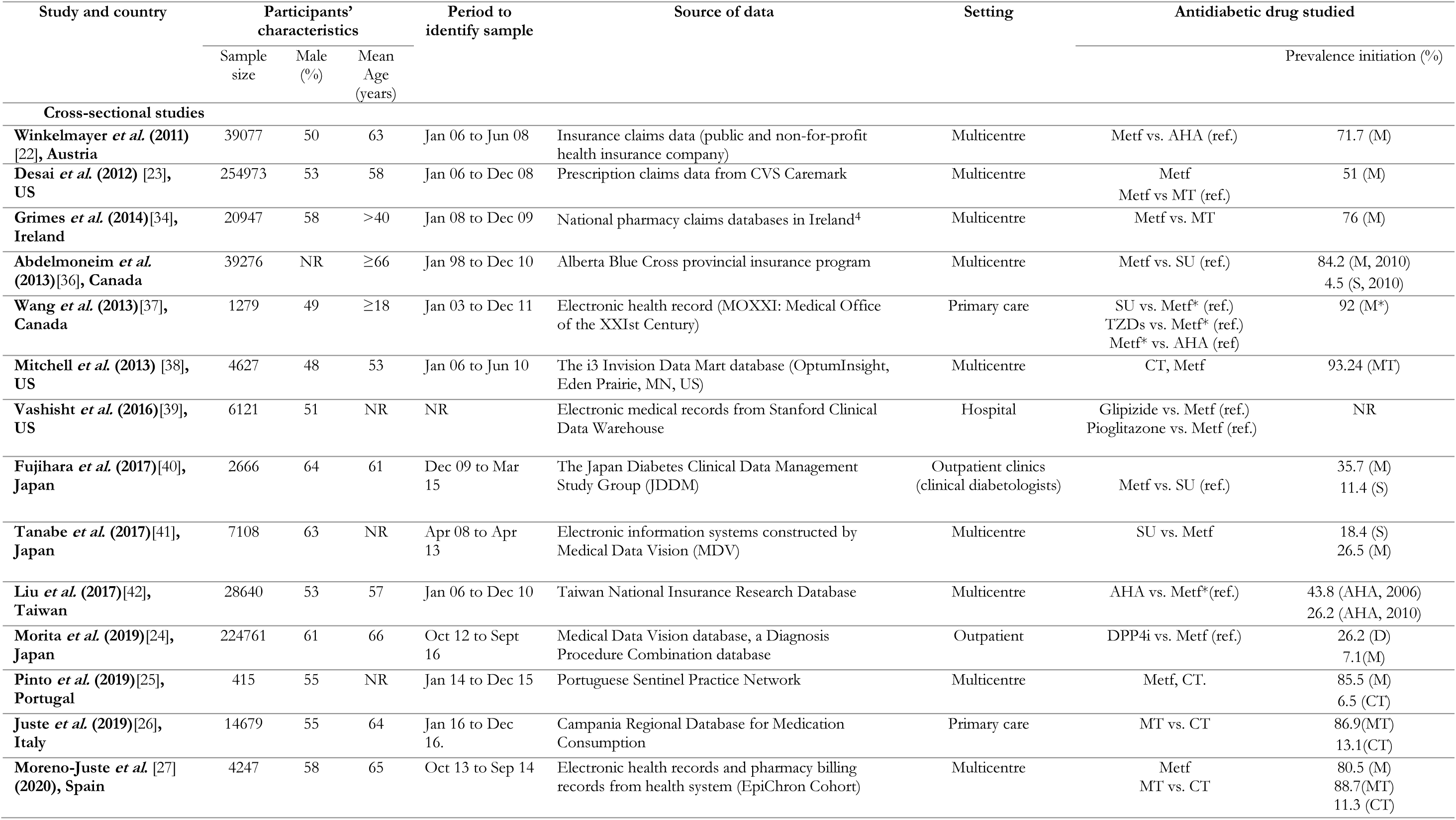
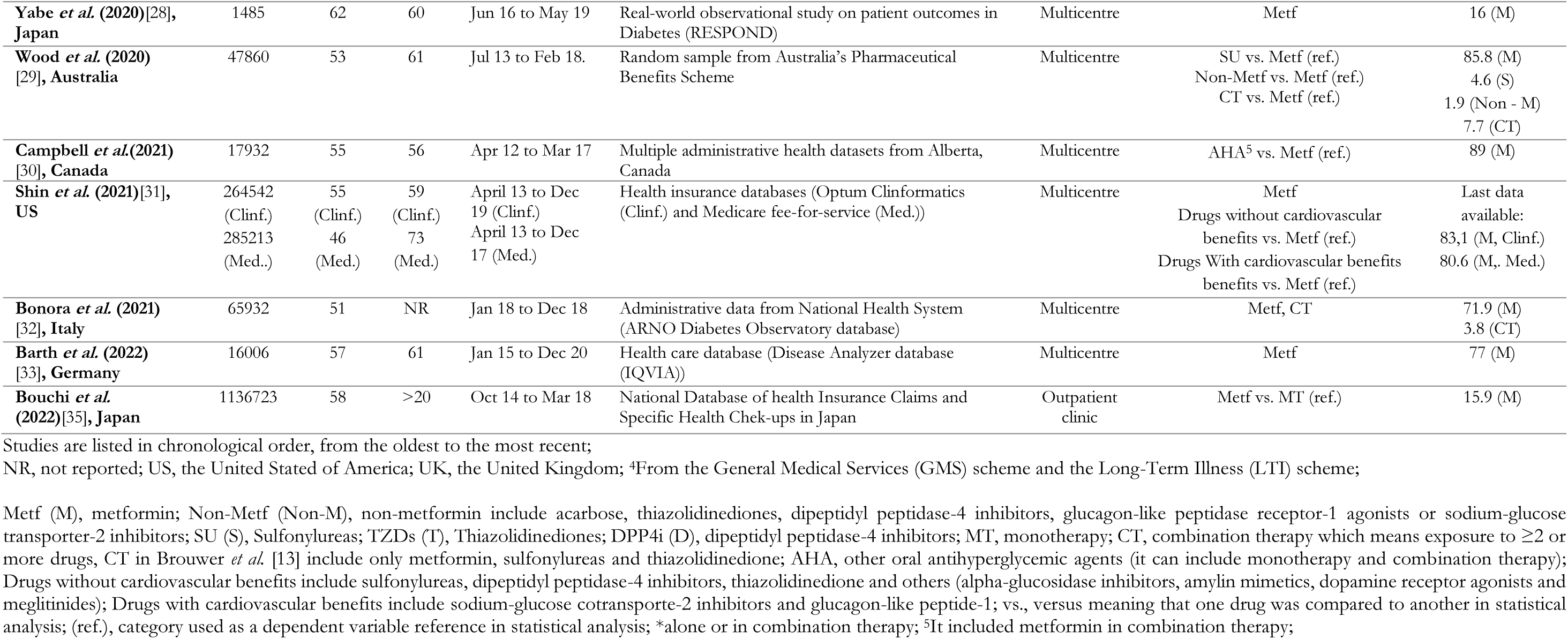
Characteristics of the cross-sectional studies which were eligible for inclusion.

Twenty studies employed data from multicentre settings [14, 16, 17, 19, 20, 22, 23, 25, 27–34, 36, 38, 41, 42] and six studies from primary care or general practitioners [13, 15, 18, 21, 26, 37]. All studies relied on secondary data, defined as data collected by others for purposes different from the objectives of the research, such as medical records and healthcare billing files[43].

Twenty-eight studies [13–20, 22–25, 27–42] explored factors associated with metformin initiation. Nineteen [13, 16–19, 22–24, 29–31, 34–37, 39–42] of these studies employed statistical models that examined factors linked to metformin initiation in direct comparison to initiation of other antidiabetic drug(s). Sulfonylureas group were the most frequently compared drugs, accounting for 36.7% (n=11) of these studies [13, 16–19, 29, 36, 37, 39–41]. Eight studies [13, 21, 25–27, 29, 32, 38] examined factors influencing combination therapy initiation, and 6 studies [13, 25, 27, 29, 32, 38] analysed metformin and combination therapy in the same study.

### Data extracted and analysed

The 105 variables extracted from the thirty studies are categorized into four main groups of factors: physician (Table 4), healthcare system (Table 5), patient (Tables 6, 7 and 8), and disease factors (Table 9). The most extensive group of factors belongs to patient factors, with five subgroups of factors recognised: sociodemographic, lifestyle and metabolic, cardiovascular, renal and other clinical factors. Sixty-five variables (62%) were evaluated by one study, and 11 (10,5%) were assessed by five or more studies.

**Table 4:**
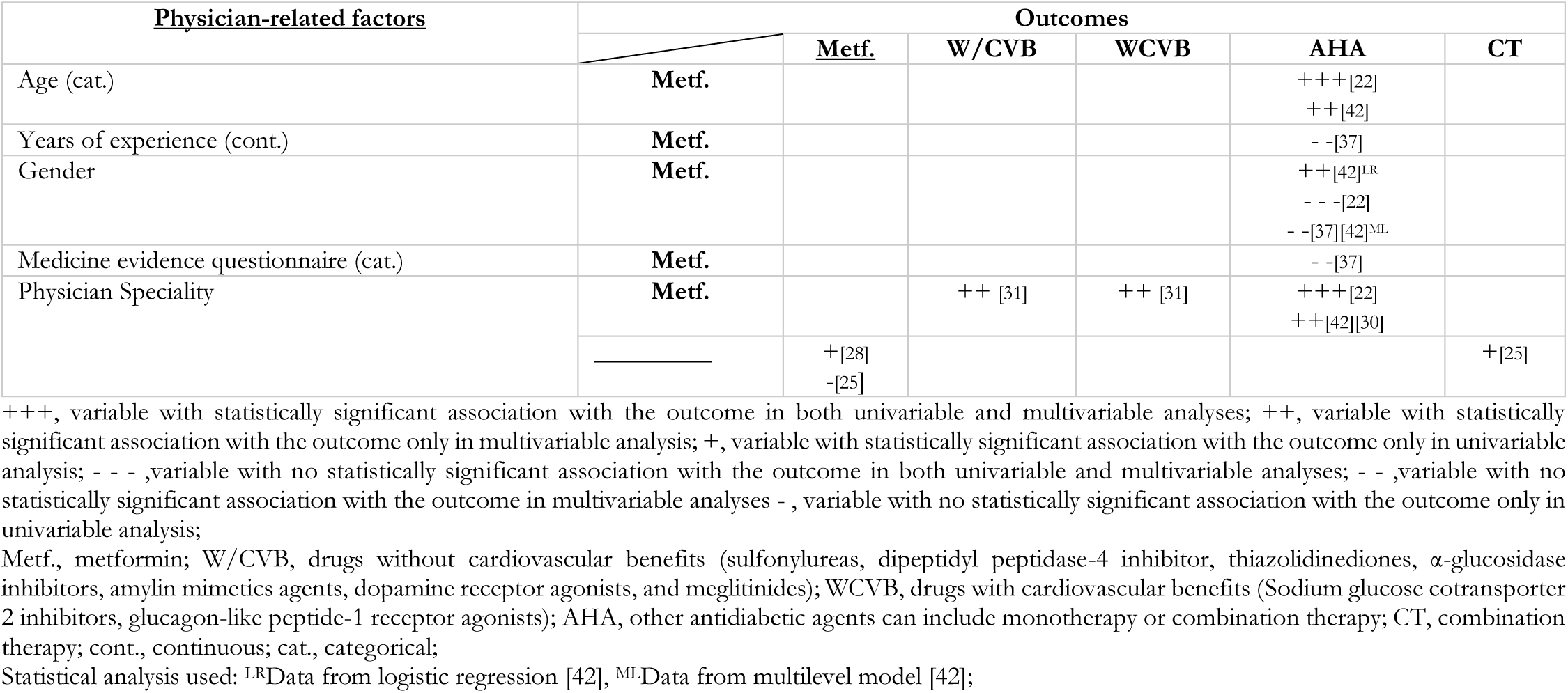
Strength of association between physician-related factors with the outcomes.

**Table 5:**
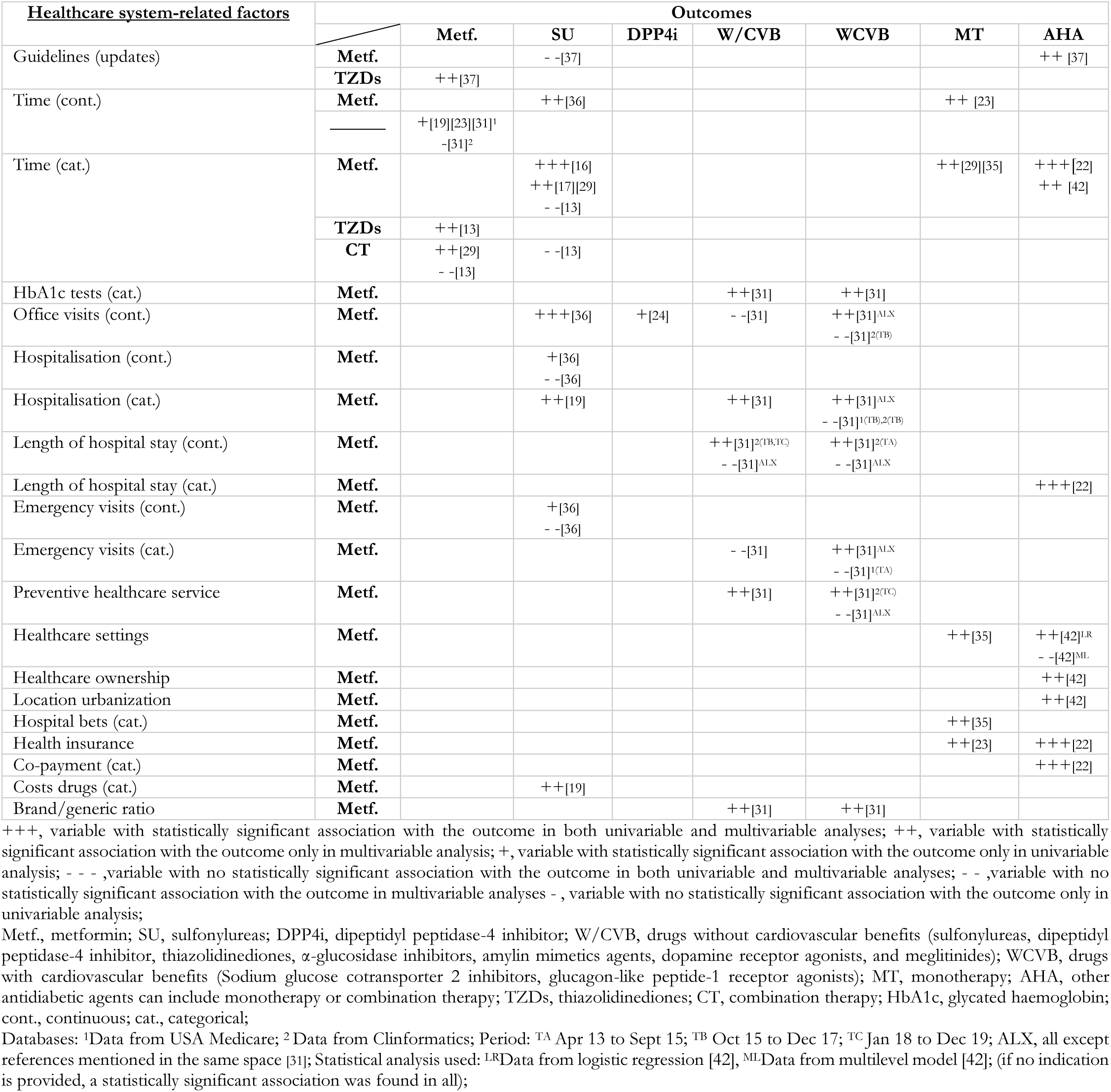
Strength of association between healthcare system-related factors with outcomes.

**Table 6:** Strength of association between patient-related factors: sociodemographic, lifestyle and metabolic with the outcomes.

| Patient-related factors Sociodemographic | Outcomes |  |  |  |  |  |  |  |
| --- | --- | --- | --- | --- | --- | --- | --- | --- |
|  |  | Metf. | SU | W/CVB | WCVB | MT | AHA | CT |
| Age (cont.) | Metf. |  | +++[36]<br>++[17][19][40]<br>+[16][41] | ++[31] | ++[31] |  | ++[30] |  |
|  | DPP4i | + [24] |  |  |  |  |  |  |
|  | CT |  |  |  |  | + [26][27] |  |  |
| Age (cat.) | Metf. |  | +++ [16]<br>++ [18][13][29] |  |  | ++ [23][34][29] | +++ [22]<br>++ [37][42] |  |
|  | TZDs | ++ [13] |  |  |  |  |  |  |
|  | CT | ++ [13][29] | - - [13] |  |  |  |  |  |
|  |  | + [14][15][20] |  |  |  |  |  |  |
| Age (cat.) and health insurance | Metf. |  |  |  |  | ++ [35] |  |  |
| Gender | Metf. |  | +++ [16]<br>++ [19]<br>- - - [36]<br>- - [17][13][40][29] | ++ [31]<sup>ALX</sup><br>- - [31]<sup>2(TC)</sup> | ++ [31]<sup>ALX</sup><br>- - [31]<sup>1(TA,TB)</sup> | ++ [23][34][35]<br>- - [29] | +++ [22]<br>++ [42]<br>- - [37][30] |  |
|  | TZDs | - - [13] |  |  |  |  |  |  |
|  | DPP4i | + [24] |  |  |  |  |  |  |
|  | CT | ++ [13][29] | - - [13] |  |  | + [26]<br>- [27] |  |  |
|  | — | - [32] |  |  |  |  |  | + [32]<sup>3</sup><br>- [32]<sup>3</sup> |
| Race/Ethnicity | Metf. |  | +++ [16]<br>++ [19][13] | ++ [31]<sup>1(TA,TB)</sup> | ++ [31]<sup>1(TA,TB)</sup> |  |  |  |
|  | TZDs | - - [13] |  |  |  |  |  |  |
|  | CT | - - [13] | - - [13] |  |  |  |  |  |
| Socioeconomic status | Metf. |  | ++ [19]<br>+ [16] |  |  | ++ [23] | ++ [42]<sup>LR</sup><br>- - [42]<sup>ML</sup> |  |
|  | CT |  |  |  |  | - [27] |  |  |
| Doctor or has a doctor in family. | Metf. |  |  |  |  |  | ++ [42]<sup>LR</sup><br>- - [42]<sup>ML</sup> |  |
| Geographic region | Metf. |  | ++ [19] | ++ [31] | ++ [31] |  |  |  |
|  | CT |  |  |  |  | + [26][27] |  |  |
| Immigrant status | Metf. |  |  |  |  | + [27] |  |  |
| Lifestyle and metabolic |  |  |  |  |  |  |  |  |
| BMI (cont.) | Metf. |  | ++ [17][40]<br>+ [16] |  |  |  |  |  |
|  | DPP4i | + [24] |  |  |  |  |  |  |
| BMI (cat.) | Metf. |  | ++ [18] |  |  |  |  |  |
| Obesity or overweight | Metf. |  |  | ++ [31] | ++ [31] |  |  |  |
| Smoker (cat.) | Metf. |  |  | ++ [31]<sup>2(TA,TB)</sup><br>- - [31]<sup>ALX</sup> | ++ [31]<sup>ALX</sup><br>- - [31]<sup>1(TB),2(TC)</sup> |  |  |  |
| Ex-smoker (cat.) | Metf. |  | + [16]<br>- - [16] |  |  |  |  |  |
| Current smoker (cat.) | Metf. |  | - - - [16]<br>- - [17] |  |  |  |  |  |
| Substance abuse (cat.) | Metf. |  |  | ++ [31]<sup>2(TA)</sup><br>- - [31]<sup>ALX</sup> | ++ [31]<sup>ALX</sup><br>- - [31]<sup>1(TA,TB)</sup> |  |  |  |
| Liver disease | Metf. |  | +++ [36]<br>++ [18] |  |  |  |  |  |
|  | DPP4i | - [24] |  |  |  |  |  |  |
| Hyperlipidaemia | Metf. |  | - [16] | ++ [31]<sup>ALX</sup><br>- - [31]<sup>1(TA),2(TA)</sup> | ++ [31] |  |  |  |
| Dyslipidaemia | Metf. |  | ++ [29] |  |  | ++ [29] |  | ++ [29] |
| HDL (cont.) | Metf. |  | - [16] |  |  |  |  |  |
| LDL (cont.) | Metf. |  | + [16] |  |  |  |  |  |
| Lipid-lowering meds | Metf. |  | +++ [36]<br>- - [17] |  |  | ++ [35] |  |  |
| Statin use | Metf. |  |  | ++ [31] | ++ [31] |  |  |  |

+++, variable with statistically significant association with the outcome in both univariable and multivariable analyses; ++, variable with statistically significant association with the outcome only in multivariable analysis; +, variable with statistically significant association with the outcome only in univariable analysis; --, variable with no statistically significant association with the outcome in both univariable and multivariable analyses; --, variable with no statistically significant association with the outcome in multivariable analyses; -, variable with no statistically significant association with the outcome only in univariable analysis;
Metf., metformin; SU, sulfonylureas; W/CVB, drugs without cardiovascular benefits (sulfonylureas, dipeptidyl peptidase-4 inhibitor, thiazolidinediones, $\alpha$ -glucosidase inhibitors, amylin mimetics agents, dopamine receptor agonists, and meglitinides); WCVB, drugs with cardiovascular benefits (Sodium glucose cotransporter 2 inhibitors, glucagon-like peptide-1 receptor agonists); MT, monotherapy; AHA, other antidiabetic agents can include monotherapy or combination therapy; CT, combination therapy; DPP4i, dipeptidyl peptidase-4 inhibitor; TZDs, thiazolidinediones; BMI, body mass index; HDL, high-density lipoprotein; LDL, low-density lipoprotein; cont., continuous; cat., categorical; Databases: <sup>1</sup>Data from USA Medicare; <sup>2</sup>Data from Clinformatics; Period: <sup>TA</sup> Apr 13 to Sept 15; <sup>TB</sup> Oct 15 to Dec 17; <sup>TC</sup> Jan 18 to Dec 19; ALX, all except references mentioned in the same space [31]; Statistical analysis used.: <sup>LR</sup>Data from logistic regression [42], <sup>ML</sup>Data from multilevel model [42]; (if no indication is provided, a statistically significant association was found in all);
<sup>3</sup>Combination therapy between metformin and DPP4i, metformin and SGLT2i, metformin and TZDs (pioglitazone) showed statistical significance, and combination therapy between metformin and sulfonylureas, TZDs (pioglitazone) and DPP4i (alogliptin) did not show statistical significance;

**Table 7:**
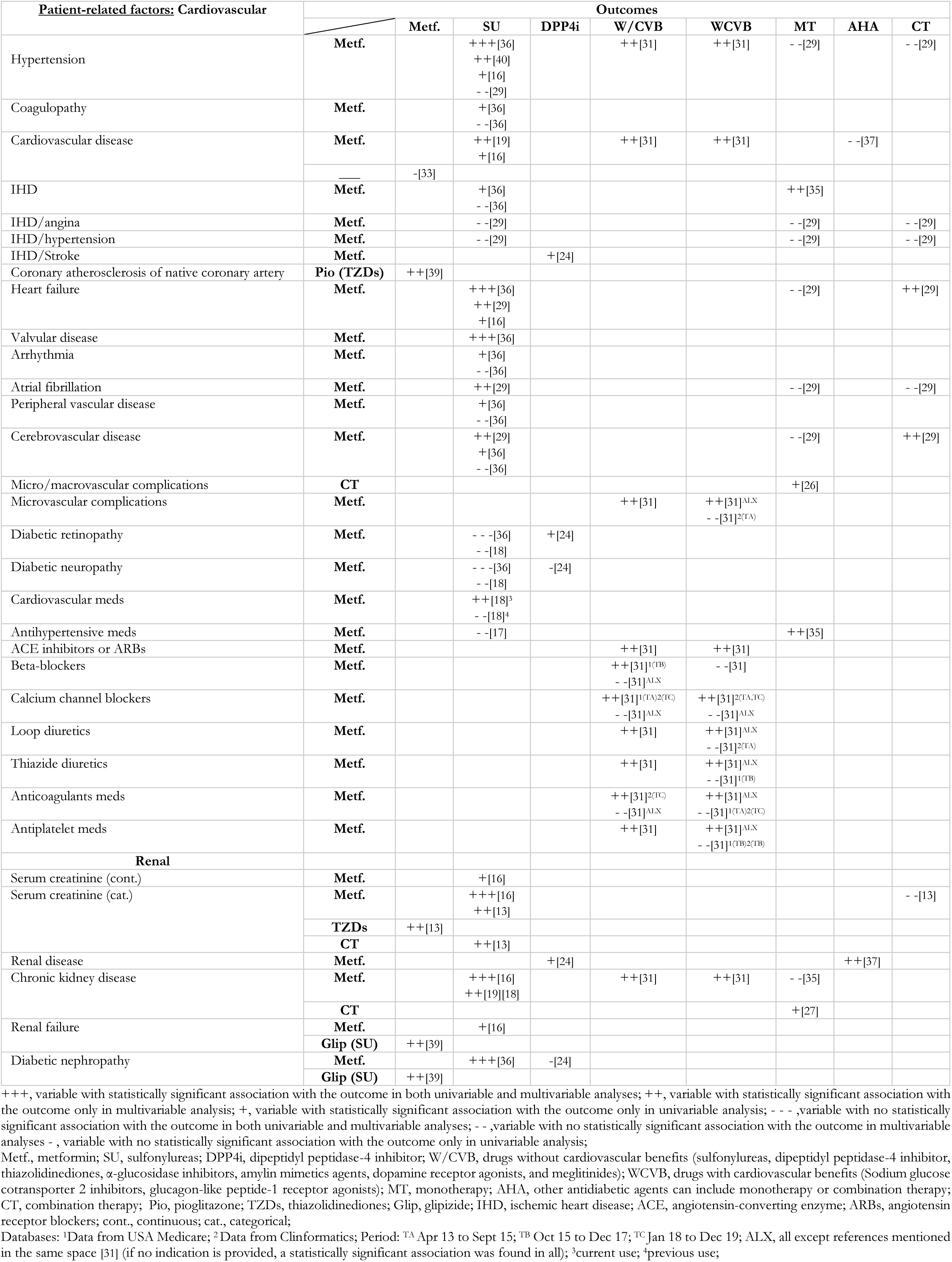
Strength of association between patient-related factors: cardiovascular and renal with the outcomes.

**Table 8:** Strength of association between patient-related factors: other clinical with the outcomes.

| Patient-related factors: Other clinical | Outcomes |  |  |  |  |  |  |  |  |
| --- | --- | --- | --- | --- | --- | --- | --- | --- | --- |
|  |  | Metf. | SU | DPP4i | W/CVB | WCVB | MT | AHA | CT |
| Esophageal varices, without bleeding, in disease classified elsewhere | Glip (SU) | ++[39] |  |  |  |  |  |  |  |
| Fluid and electrolyte disorder | Metf. |  | + [36]<br>- -[36] |  |  |  |  |  |  |
| Paracetamol | Pio (TZDs) | ++[39] |  |  |  |  |  |  |  |
| COPD | Metf. |  | + [36]<br>- -[36] |  | ++[31]<sup>1(TA)2(TA)</sup><br>- -[31]<sup>ALX</sup> | ++[31]<sup>1(TC)</sup><br>- -[31]<sup>ALX</sup> |  |  |  |
| Pulmonary collapse | Pio (TZDs) | ++[39] |  |  |  |  |  |  |  |
| Dementia | Metf. |  | ++[18] |  |  |  |  |  |  |
| Depression | Metf. |  | ++[29]<br>+[16][36]<br>- -[36] |  |  |  | - -[29] |  | ++[29] |
| Neuro-psychiatric meds | CT |  |  |  |  |  | + [26] |  |  |
| Antipsychotic meds | Metf. |  | - -[18]<sup>3,4</sup> |  |  |  |  |  |  |
| Cancer | Metf. |  | +++[36] |  |  |  |  |  |  |
| Lymphoma | Metf. |  | + [36]<br>- -[36] |  |  |  |  |  |  |
| Hypothyroidism | Metf. |  | - - -[36] |  |  |  |  |  |  |
| Rheumatoid arthritis | Metf. |  | + [36]<br>- -[36] |  |  |  |  |  |  |
| Immune modulators/suppressants | Metf. |  | ++[18]<sup>3</sup><br>- -[18]<sup>4</sup> |  |  |  |  |  |  |
| Oral corticosteroids | Metf. |  | ++[18]<sup>3</sup><br>- -[18]<sup>4</sup> |  |  |  |  |  |  |
| Tacrolimus | Glip (SU) | ++[39] |  |  |  |  |  |  |  |
| Cefepime | Glip (SU) | ++[39] |  |  |  |  |  |  |  |
| Medication use (cont.) | Metf. |  | - -[19] |  |  |  | ++[23] |  |  |
|  | CT |  |  |  |  |  | + [27] |  |  |
| Medication use (cat.) | Metf. |  |  |  |  |  |  | +++[22] |  |
|  | CT |  |  |  |  |  | + [26] |  |  |
| Comorbidities (cont.) | Metf. |  | ++[19] |  |  |  |  |  |  |
|  | CT |  |  |  |  |  | + [27] |  |  |
| Comorbidities (cat.) | Metf. |  | ++[29] |  |  |  | ++[29] | ++[30] | ++[29] |
| Quan Score (cont.) | Metf. |  | + [16] |  |  |  |  |  |  |
| Rx-Risk comorbidity index (cont.) | CT |  |  |  |  |  | + [26] |  |  |

+++ , variable with statistically significant association with the outcome in both univariable and multivariable analyses; ++ , variable with statistically significant association with the outcome only in multivariable analysis; + , variable with statistically significant association with the outcome only in univariable analysis; - - - , variable with no statistically significant association with the outcome in both univariable and multivariable analyses; - - , variable with no statistically significant association with the outcome in multivariable analyses - , variable with no statistically significant association with the outcome only in univariable analysis;
Metf., metformin; SU, sulfonylureas; DPP4i, dipeptidyl peptidase-4 inhibitor; W/CVB, drugs without cardiovascular benefits (sulfonylureas, dipeptidyl peptidase-4 inhibitor, thiazolidinediones, $\alpha$ -glucosidase inhibitors, amylin mimetics agents, dopamine receptor agonists, and meglitinides); WCVB, drugs with cardiovascular benefits (Sodium glucose cotransporter 2 inhibitors, glucagon-like peptide-1 receptor agonists); MT, monotherapy; AHA, other antidiabetic agents can include monotherapy or combination therapy; CT, combination therapy; Glip, glipizide; Pio, pioglitazone; TZDs, thiazolidinediones; COPD, chronic obstructive pulmonary disease; cont., continuous; cat., categorical;
Databases: <sup>1</sup>Data from USA Medicare; <sup>2</sup>Data from Clinformatics; Period: <sup>TA</sup> Apr 13 to Sept 15; <sup>TB</sup> Oct 15 to Dec 17; <sup>TC</sup> Jan 18 to Dec 19; ALX, all except references mentioned in the same space [31] (if no indication is provided, a statistically significant association was found in all); <sup>3</sup>current use; <sup>4</sup>previous use;

**Table 9:**
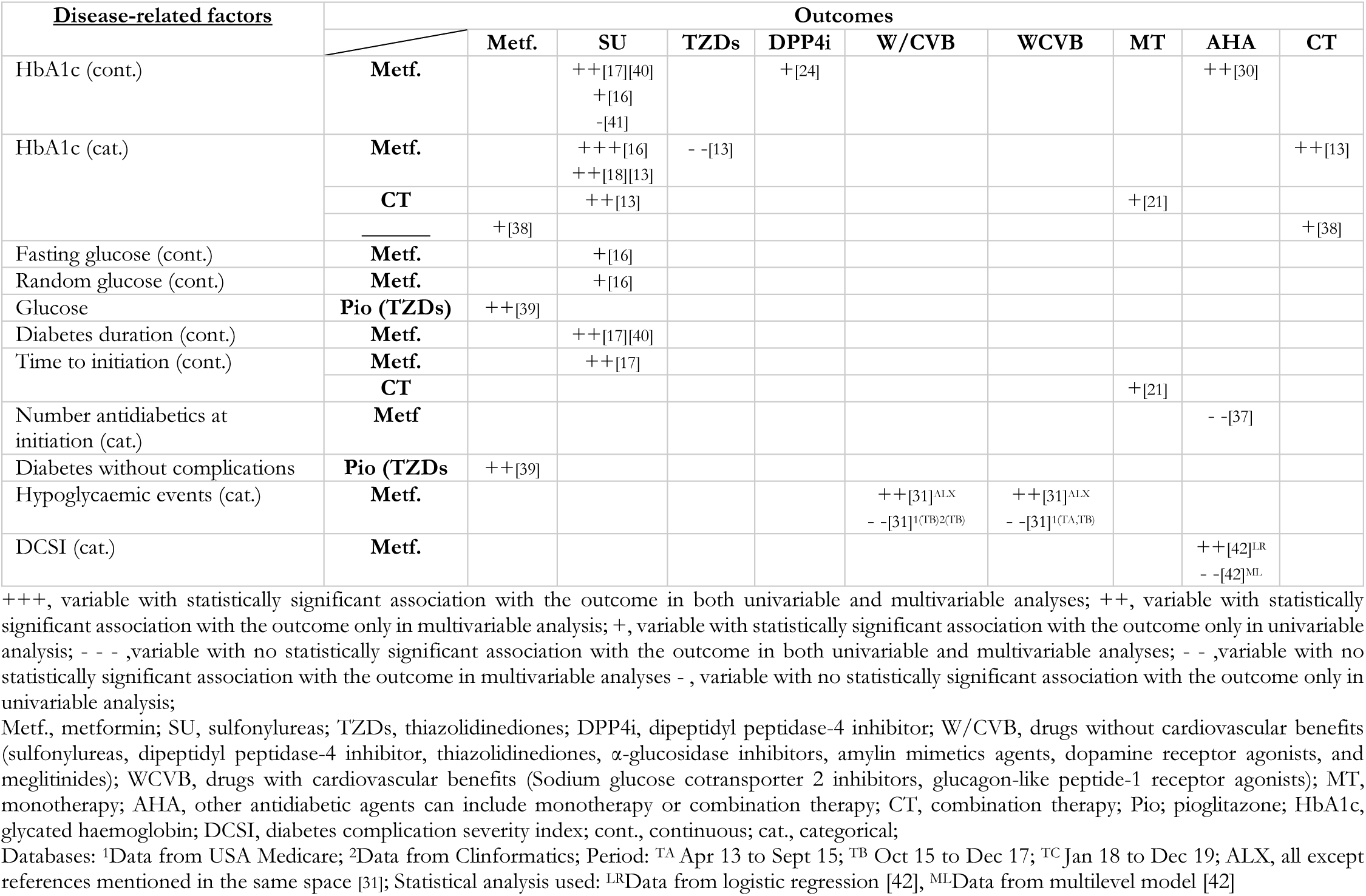
Strength of association between disease-related factors with the outcomes.

It is also important to highlight that one study [31] showed data analysis not as a single block, but for different periods and two databases, and another [42] employed two statistical analyses: multivariable logistic regression and multilevel linear model.

### Physician-related factors

Table 4 shows that physician age presented a statistical association with the prescription profile [22, 42], with older physicians prescribing non-metformin treatments more frequently than metformin. Three studies [22, 37, 42] reported a non-association between gender and initial therapy choice. Only Liu *et al*. [42] found that male physicians had a higher chance of prescribing non-metformin than female physicians in logistic regression. Physician speciality was the most assessed variable in this category, with six studies [22, 25, 28, 30, 31, 42] showing a statistically significant association. Campbell *et al*.[30] stated that specialists were more likely to prescribe metformin in combination or other drug therapies instead of metformin alone compared to general practitioners (GPs). Pinto *et al.* [25] also reported that GPs had a lower prevalence of prescribed combined therapy than other specialists (4.2% vs 33.3%, p-value <0.001). However, no significant differences were found in the prescription rates of metformin alone or in combination between GPs and other specialists. Another study [42] presented that GPs had a higher chance of prescribing non-metformin than metformin compared to endocrinologists. In contrast, Shin *et al.*[31] observed that individuals visiting endocrinologists had a lower chance of initiating metformin than other drugs independently of their cardiovascular benefits; the opposite was found when visiting internists. Finally, the two additional variables, years of experience and the medical evidence questionnaire, assessed by Wang *et al.* [37] presented a non-statistically significant association.

### Healthcare system-related factors

Eleven studies [13, 16, 17, 19, 22, 23, 29, 31, 35, 36, 42] demonstrated a statistically significant association between more recent time periods and the prescription profile. However, in two studies, this depended on the analysed database [31] or medicine and the respective comparison [13] to report statistical significance. Studies reported that the most recent periods were positively associated with metformin initiation [19, 31], and also when compared with sulfonylureas [16, 17, 29, 36], other monotherapies [23, 35], other antidiabetic agents (eventually in monotherapy or combination therapy) [22, 42] or combination therapy [29]. Additionally, Wang *et al*. [37] studied how primary care physicians responded to a change in the Canadian Diabetes Association Guidelines, which significantly increased metformin initiation as a first line, except when compared to sulfonylureas.

One cross-sectional study [31] reported that individuals with three or more HbA1c tests within 365 days before the index date had higher odds of initiating any other medicine than metformin, irrespective of their cardiovascular benefits. Additionally, variables such as the number of office visits [24, 36], hospitalisations [19, 36], emergency visits [36], and the length of stay [22] were statistically significantly associated with lower odds of metformin prescriptions. However, Abdelmoneim *et al*. [36] and Shin *et al.* [31] reported no statistically significant association with some of these variables (see Table 5).

Two studies [22, 23] reported an association between the health insurance and the initial therapy choice. Regarding the co-payment and cost of drugs, one study [22] indicated that individuals with co-payment waiver had a lower chance of starting metformin than sulfonylureas, the same that Li [19] found for individuals in the top 10% of prescription drug expenses under Medicare Part D. Medicare is a health program for individuals aged 65 and over and younger individuals with disabilities, with Medicare Part D being an optional plan that covers drug costs[44]. Shin *et al*. [31] stated that individuals with more brand-name experience also had a lower chance of starting metformin than drugs independently of their cardiovascular benefits.

### Patient-related factors: Sociodemographic

Twenty-two studies [13–20, 22–24, 26, 27, 29–31, 34, 36, 37, 40–42] evaluated the association between age and initial therapy choice. Metformin prescription was associated with younger individuals compared to sulfonylureas [13, 16–19, 29, 36, 40, 41], other antidiabetic agents (including monotherapy (MT) and eventually MT and combination therapy (AHA)) [22, 29, 30, 34, 37, 42], thiazolidinediones [13], DPP4i [24], and drugs without cardiovascular benefits [31]. Even when the interaction between age and type of insurance was studied [35], metformin prescription decreased with advancing age compared to other monotherapies. On the other hand, when metformin is compared with drugs with cardiovascular benefits, the chance of prescribing increases with age [31]. Two studies [13, 29] found a significant but inverse association between age and combination therapy. The other two studies [26, 27] indicated that younger people were more prevalent in combination therapy than monotherapy. Moreover, one study [13] found no association between age and combination therapy compared to sulfonylureas.

Nineteen studies tested the association between gender and prescription profile [13, 16, 17, 19, 22–24, 26, 27, 29–32, 34–37, 40, 42]. Nine studies [16, 19, 22–24, 26, 34, 35, 42] showed a statistically significant association and six none[17, 27, 30, 32, 37, 40]. In the remaining four studies [13, 29, 31, 32] several analysis were conducted within each study. This led to the report of statistically significant associations depending on the prescribing profile evaluated (i.e., the same study analysed different prescribing profiles with gender) or the database analysed (i.e., the same study analysed two different databases). The statistical associations found that females were more likely to initiate metformin than men compared to sulfonylureas [16, 19], other monotherapies [34, 35], other antidiabetic agents [42] or DPP4i [24]. On the other hand, Winkelmayer *et al.* [22] identified a significant association, though its direction varied between univariate and multivariate analyses, and Desai *et al.*[23] noted that men had a higher chance of starting metformin than other monotherapies.

Regarding race/ethnicity, black individuals showed a lower chance of starting metformin than sulfonylureas compared to white individuals [13, 16, 19]. White individuals also showed more chance of starting drugs with cardiovascular benefits than metformin compared to non-white [31]. However, there was no statistically significant association between race/ethnicity and combination therapy compared to metformin or sulfonylureas [13].

Individuals with lower socioeconomic status were associated with more frequent non-metformin prescriptions [23, 42] or sulfonylurea prescriptions [16, 19], compared to metformin. Conversely, Liu *et al.* [42] also found the same for a higher socioeconomic status category, and statistical significance was lost in the multilevel linear model.

### Patient-related factors: Lifestyle and metabolic

Nine studies [16–18, 24, 29, 31, 35, 36, 40] evaluated at least one lifestyle or metabolic variable (Table 6). A higher body mass index (BMI) was significantly associated with an increased chance of starting metformin compared to sulfonylureas [16–18, 40], DPP4i [24], or drugs without cardiovascular benefits[31]. However, when compared to drugs with cardiovascular benefits, the chance of prescribing metformin decreased with a higher BMI [31]. Liver disease was associated with a decreased chance of starting metformin compared to sulfonylureas [18, 36]. However, no statistical significance was found when compared to DPP4i. Individuals with dyslipidaemia showed higher odds of initiating combination therapy than metformin [29]. For other variables in this group, either no statistical significance was found, or the direction of associations was inconclusive.

### Patient-related factors: Cardiovascular

The impact of twenty-seven cardiovascular variables (the most representative group of variables) was assessed in fourteen studies [16–19, 24, 26, 29, 31, 33, 35–37, 39, 40]. Hypertension was evaluated with initial therapy choice in five studies [16, 29, 31, 36, 40]. Three studies [16, 36, 40] showed statistical significance, but with conflicting results. Abdelmoneim *et al.* [36] found that hypertension increased the odds of starting metformin compared to sulfonylureas, while Fujihara *et al*. [40] reported the opposite. Shin *et al.* [31] also linked hypertension to a decreased chance of starting metformin compared to drugs with or without cardiovascular benefits. However, Shin *et al.* [31] found that the use of angiotensin-converting enzyme inhibitors and angiotensin receptor blockers (antihypertensive drugs) increased the odds of initiating metformin. No significant association was found between hypertension and combination therapy compared to metformin [29].

Two studies [16, 19] reported that cardiovascular disease was negatively associated with metformin initiation compared to sulfonylureas. Similarly, Shin *et al.* [31]found the same when comparing metformin to drugs with or without cardiovascular benefits. No statistically significant association was found by Wang *et al*. [37].

Three studies [16, 29, 36] reported that heart failure was negatively associated with metformin compared to sulfonylureas. Additionally, Wood *et al.* [29] reported that individuals with heart failure had higher odds of initiating combination therapy rather than metformin. Similarly, cerebrovascular disease decreased the odds of initiating metformin compared to sulfonylureas or combination therapy, although no significant association was found when comparing metformin to non-metformin therapies for either heart failure or cerebrovascular disease [29]. The valvular disease kept the tendency reported, and it was associated with a decreased chance of initiating metformin compared to sulfonylureas [36].

### Patient-related factors: Renal

Eleven studies[13, 16, 18, 19, 24, 27, 31, 35–37, 39] evaluated renal-related variables. It is unanimously reported that renal problems, such as elevated serum creatinine, renal disease, chronic renal disease (CKD), renal failure and diabetes-related nephropathy decreased the chance of metformin initiation [13, 16, 18, 19, 24, 31, 35–37, 39]. For example, Raebel *et al*.[16] showed that individuals with serum creatine range of 1.4 to ≤2 mg/dl compared to individuals with <1.4mg/dl (reference) had a relative risk of 2.21 (95% CI, 2.05-2.39) of starting sulfonylureas compared to metformin and Wang *et al*. [37] also showed that individuals with renal disease had lower odds (OR 0.14; 95% CI, 0.05-0.40) of starting metformin than other antidiabetic agents.

Regarding combination therapy, Brouwer *et al*. [13] reported that high serum creatinine decreased the chance of beginning combination therapy compared to sulfonylureas, and another study [27] stated a lower prescription of combination therapy compared to monotherapies such as sulfonylureas or DPP4i among individuals with CKD [27].

### Patient-related factors: Other clinical

Twenty-one other clinical variables were assessed with the prescription profile in twelve studies [16, 18, 19, 22, 23, 26, 27, 29–31, 36, 39]. Of these, 81% (17) of the variables were assessed by one study. Dementia was negatively associated with metformin initiation compared to sulfonylureas [18]. Wood *et al.* [29] reported that depression was positively associated with metformin initiation compared to sulfonylureas, which aligned with Raebel *et al.* [16] findings. However, these associations differed in direction from the univariable analysis results reported by Abdelmoneim *et al*.[36]. Wood *et al*. [29] also reported that depression was positively associated with metformin initiation compared with combination therapy, and statistical significance was not found comparing non-metformin monotherapies with metformin. Juste *et al*. [26] reported that the prevalence of neuro-psychiatric medication was higher among monotherapy initiators than among combination therapy initiators.

Regarding medication use, Desai *et al.* [23] reported that the chances of starting metformin than other monotherapies decreased for each additional prescription. However, Li [19] found no statistical association, and Winkelmayer *et al*. [22] observed that the odds of initiating metformin increased with a higher number of therapeutic class prescriptions compared to other antidiabetic agents. However, this pattern shifted in the univariable analysis, where individuals taking ≥9 medications had reduced odds of initiating metformin compared to those taking none. Two studies[26, 27] stated that the prevalence another type of medication was higher among monotherapy initiators than among combination therapy initiators.

Wood *et al*. [29] presented that individuals with one to three comorbidities had a lower chance of starting sulfonylureas than metformin compared to those with no comorbidities. No statistically significant association was found when comparing four or more comorbidities to zero. Additionally, individuals with one to six comorbidities had lower odds of initiating non-metformin monotherapy compared to metformin, though this was not statistically significant when comparing seven or more comorbidities to zero. Nevertheless, compared to zero, one or more comorbidities reduced the odds of initiating combination therapy rather than metformin. Campbell *et al.* [30] also reported that individuals with one or more comorbidities were less likely to start metformin in combination therapy or other drug therapies, instead of metformin alone compared to those with no comorbidities. Additionally, Juste *et al*. [26] noted that individuals who initiated combination therapy had a lower comorbidity score compared to individuals who initiated monotherapy.

### Disease-related factors

Data was extracted from fourteen studies [13, 16–18, 21, 24, 30, 31, 37–42] with ten variables collected. HbA1c was the most studied variable, being addressed in ten studies [13, 16–18, 21, 24, 30, 38, 40, 41]. Five studies [13, 16–18, 40] stated that higher HbA1c levels were statistically associated with the initial sulfonylureas therapy instead of metformin, but one study did not find statistical significance [41]. Campbell *et al.* [30] also indicated that individuals with a higher HbA1c level were more likely to start non-metformin and combination therapy than metformin alone. On the other hand, Morita *et al.* [24] reported the opposite when comparing metformin to DPP4i, and no statistically significant association was noted when comparing metformin to thiazolidinediones [13]. Combination therapy was statistically associated with higher levels of HbA1c compared to metformin, sulfonylureas [13] or monotherapy [21].

Raebel *et al.* [16] also assessed fasting and random glucose levels and reported that both levels were higher among those who initiated sulfonylureas than those who initiated metformin in univariable analysis. Vashisht *et al.* [39] identified glucose and diabetes without complications as variables statistically associated with pioglitazone (TZD) choice instead of metformin. However, the researchers did not specify how glucose was measured, nor did they clarify whether the association with pioglitazone was positive or negative.

Shorter diabetes duration [17, 40] and earlier treatment initiation [17] were statistically associated with metformin initiation compared to sulfonylureas. Ouchi *et al.* [21] also found that combination therapy (89.62 days, SD 279.1) was prescribed significantly earlier (p<0.001) than monotherapy (190.7 days, SD 366.2).

### Quality assessment

The quality assessment results for observational cohort studies are presented in Table 10. None of the studies ensured the similarity between compared groups (exposed vs. unexposed). It was also impossible to ensure exposures measured similarly between groups and their validity and reliability as all studies relied on secondary data without detailing the method of exposure measurement. None of the studies identified potential confounders or strategies to address them. Five studies [13, 16, 17, 19, 21] raised concerns about outcome validity and reliability, as the data sources might have reflected treatment adherence (e.g., pharmacy dispensing records) rather than prescriptions. Follow-up loss was either ignored [17] or avoided through inclusion/exclusion criteria [13–16, 18–21] without strategies being expected to address it. Although five studies [13, 16–19] used multivariable analysis (appropriate statistical analysis considered), none reported statistical model assumptions, and one study [21] did not report the applied statistical analysis.

**Table 10:** Quality assessment and risk of bias of cohort studies included.

| Retrospective cohort studies | Topics assessed |  |  |  |  |  |  |  |  |  |  |
| --- | --- | --- | --- | --- | --- | --- | --- | --- | --- | --- | --- |
|  | D1 | D2 | D3 | D4 | D5 | D6 | D7 | D8 | D9 | D10 | D11 |
| Brouwer et al.[13] | Unclear | Unclear | Unclear | No | No | Unclear | Unclear | Yes | Unclear | N/A | Unclear |
| Zhang et al. [14] | Unclear | Unclear | Unclear | No | No | Yes | Yes | Yes | Unclear | N/A | No |
| Sinclair et al. [15] | Unclear | Unclear | Unclear | No | No | Yes | Yes | Yes | Unclear | N/A | No |
| Raebel et al. [16] | Unclear | Unclear | Unclear | No | No | Yes | Unclear | Yes | Unclear | N/A | Unclear |
| Geier et al. [17] | Unclear | Unclear | Unclear | No | No | Yes | Unclear | Yes | Unclear | N/A | Unclear |
| Wright [18] | Unclear | Unclear | Unclear | No | No | Yes | Yes | Yes | Unclear | N/A | Unclear |
| Li [19] | Unclear | Unclear | Unclear | No | No | Yes | Unclear | Yes | Unclear | N/A | Unclear |
| Carrillo Balam [20] | Unclear | Unclear | Unclear | No | No | Yes | Yes | Yes | Unclear | N/A | No |
| Ouchi et al. [21] | Unclear | Unclear | Unclear | No | No | Unclear | Unclear | Yes | Unclear | N/A | Unclear |

D1. Were the two groups similar and recruited from the same population? D2. Were the exposures measured similarly to assign people to both exposed and unposed groups? D3. Was the exposure measured in a valid and reliable way? D4. Were confounding factors identified? D5. Were strategies to deal with confounding factors stated? D6. Were the groups/participants free of the outcome at the start of the study (or at the moment of exposure)? D7. Were the outcomes measured in a valid and reliable way? D8. Was the follow up time reported and sufficient to be long enough for outcomes to occur? D9. Was follow up complete, and if not, were the reasons to loss to follow up described and explored? D10. Were strategies to address incomplete follow up utilized? D11. Was appropriate statistical analysis used? N/A, not applicable;

Table 11 presents the results of quality assessment for observational cross-sectional studies. Eight studies [24, 28, 30, 32, 38–41] did not provide clear inclusion criteria, and eight studies [23, 25, 32–34, 39–41] lacked information to infer the health status of the sample. It was also impossible to ensure exposures measured validity and reliability. However, four studies [23, 25, 32, 34] reported exposures without a gold standard measurement (e.g., age, sex, type of insurance), suggesting no risk of bias. Only two studies [30, 45] identified potential confounders, but their statistical models also examined associations with the outcome, raising doubts about their exclusive use for control. Therefore, only two other studies [28, 33] addressed confounder factors by matching or stratification. Twelve studies [22, 23, 26, 27, 29–31, 34, 36–38, 42] relied on pharmacy dispensing data or lacked clarity on data sources, undermining outcome validity and reliability. None used appropriate statistical analyses due to missing multivariable analysis [24–28, 32, 33, 38, 41], unreported statistical model assumptions[22, 23, 29–31, 34–37, 40, 42], or omission of p-value results[39]. Therefore, the quality assessment underscores multiple issues across the studies, pointing to a high risk of bias.

**Table 11:** Quality assessment and risk of bias of cross-sectional studies included.

| Cross-sectional studies | Topics assessed |  |  |  |  |  |  |  |
| --- | --- | --- | --- | --- | --- | --- | --- | --- |
|  | D1 | D2 | D3 | D4 | D5 | D6 | D7 | D8 |
| Winkelmayer et al.[22] | Yes | Yes | Unclear | Yes | No | No | Unclear | Unclear |
| Desai et al.[23] | Yes | No | N/A | Yes | No | No | Unclear | Unclear |
| Grimes et al. [34] | Yes | No | N/A | Yes | No | No | Unclear | Unclear |
| Abdelmoneim et al.[36] | Yes | Yes | Unclear | Yes | No | No | Unclear | Unclear |
| Wang et al. (2013)[37] | Yes | Yes | Unclear | Yes | Yes | Unclear | Unclear | Unclear |
| Mitchell et al.[38] | No | Yes | Unclear | Unclear | No | No | Unclear | No |
| Vashisht et al.[39] | No | No | Unclear | Yes | No | No | Yes | Unclear |
| Fujihara et al. [40] | No | No | Unclear | Unclear | No | No | Yes | Unclear |
| Tanabe et al. [41] | No | No | Unclear | Unclear | No | No | Yes | No |
| Liu et al. [42] | Yes | Yes | Unclear | Yes | No | No | Unclear | Unclear |
| Morita et al. [24] | No | Yes | Unclear | Yes | No | No | Yes | No |
| Pinto et al. [25] | Yes | No | N/A | Yes | No | No | Yes | No |
| Juste et al.[26] | Yes | Yes | Unclear | Yes | No | No | Unclear | No |
| Moreno-Juste et al. [27] | Yes | Yes | Unclear | Yes | No | No | Unclear | No |
| Yabe et al. [28] | No | Yes | Unclear | Unclear | No | Yes | Yes | No |
| Wood et al. [29] | Yes | Yes | Unclear | Yes | No | No | Unclear | Unclear |
| Campbell et al.[30] | No | Yes | Unclear | Yes | Yes | Unclear | Unclear | Unclear |
| Shin et al. (2021)[31] | Yes | Yes | Unclear | Yes | No | No | Unclear | Unclear |
| Bonora et al.[32] | No | No | N/A | No | No | No | Yes | No |
| Barth et al.[33] | Yes | No | Unclear | Yes | No | Yes | Yes | No |
| Bouchi et al.[35] | Yes | Yes | Unclear | Yes | Unclear | Unclear | Yes | Unclear |

D1. Were the criteria for inclusion in the sample clearly defined? D2. Were the study subjects and the setting described in detail? D3. Was the exposure measured in a valid and reliable way? D4. Were objective, standard criteria used for measurement of the condition? D5. Were confounding factors identified? D6. Were strategies to deal with confounding factors stated? D7. Were the outcomes measured in a valid and reliable way? D8. Was appropriate statistical analysis used? N/A, not applicable;

## Discussion

This systematic review identified 30 studies exploring factors influencing first-line treatment decisions in T2DM, focusing on metformin and combination therapy. Although clinical decision-making can be inherently complex, the identification of key factors serves two main purposes: first, it enables the alignment of clinical practice with evidence-based guidelines by addressing gaps in knowledge and practice patterns; second, it aids in tailoring treatment decisions to individual characteristics, thereby enhancing the quality of care and improving health outcomes.

The prevalence of the two initial therapies analysed varied widely across studies, with the greatest variation observed in metformin initiation. For instance, Morita *et al*. [24] reported that 7.1% of their sample started metformin monotherapy, while Campbell *et al*. [30] found that 89% of participants initiated metformin monotherapy. This discrepancy may be attributed to differences in national clinical guidelines: Canadian guidelines recommend metformin as the first-line treatment unless contraindicated [46], while Japanese guidelines do not specify a preferred drug for initiation [47]. This highlights how variations in regional clinical recommendations can significantly impact treatment patterns and decision-making processes.

One hundred and five variables were evaluated as potential predictive factors influencing initial therapy decisions, with only 9.5% (10) showing no association with initial therapy choice. Among these, 25 variables were assessed with combination therapy, with 18 (72%) evaluated in just one study. Notably, only 11 of the 105 variables were assessed in five or more studies, indicating limited replication and, consequently, reduced robustness for most findings.

Age and gender were the most frequently assessed variables, reflecting the accessibility of demographic data. On the other hand, physician-related factors were the group with the least variables evaluated, perhaps due to challenges in extracting this information from secondary data sources. In contrast, the subgroup of cardiovascular factors included the highest number of assessed variables, which may reflect researchers’ interest in studying these factors and the emphasis of guidelines on cardiovascular diseases in individuals with T2DM [48, 49]. Furthermore, among the twenty-one variables categorized as other clinical factors, doubts arise about their clinical basis for assessing their association with the initial therapy choice, potentially indicating that their availability is the primary reason for their assessment.

Interestingly, while physician age was associated with initial therapy choice, years of experience were not, yielding contradictory results since age is typically linked to years of experience. Moreover, a survey study [50] found that years of experience influenced the factors considered when selecting first-line treatment, and a chart review study [51] revealed that more experienced physicians were less likely to follow guidelines. These findings highlight the complexity of physician-related factors and their interplay with clinical decision-making. Additionally, regarding associations found with physician specialities, it is important to consider that access to these specialities is strongly influenced by the patient’s health status and the severity of the disease [52]. Healthcare utilization is also significantly affected by the patient’s socioeconomic status[53], and both socioeconomic status and health insurance were associated with initial therapy choice.

Patient-related factors, particularly age, also play a significant role in influencing therapy decisions, with metformin consistently linked to a younger age. A survey study [54] also reported that reasons cited by physicians to avoid initial dual therapy were often associated with patient age. Additionally, metformin initiation was negatively associated with several cardiovascular conditions and healthcare utilization, indicating a tendency to avoid metformin in individuals with poorer health status. This trend is surprising given its well-established safety profile [55] and its benefits for various conditions [56].

Concerns about these findings are heightened when comparing metformin to sulfonylureas, which are associated with an increased risk of hypoglycaemia [57]. Guidelines recommend a conservative approach to sulfonylureas, particularly in older individuals [4, 7, 58], and scientific literature highlights their potential harm in those at high risk for cardiovascular disease [59]. The preference for sulfonylureas over metformin at high HbA1c levels is not also supported by guideline recommendations [4] or scientific literature [60, 61].

On the other hand, renal-related factors present a valid reason for avoiding metformin, as it should not be used in individuals with an estimated glomerular filtration rate <30 ml/min per 1.73 m² [62]. This may explain why studies report that physicians are cautious about prescribing it to individuals with renal problems. Similarly, for BMI, the findings also align with guideline recommendations [4, 7].

The positive association between high HbA1c levels and combination therapy is consistent with recommendations, particularly the consensus report from the ADA and EASD [4], which advocates for considering initial combination therapy in individuals with elevated HbA1c at diagnosis. Additionally, the tendency to avoid combination therapy in individuals with a high number of comorbidities aligns with Ismail-Beigi *et al.* [63], who highlighted the importance of less intensive treatment for those with multiple or severe comorbidities.

While these findings are noteworthy, it is important to recognize the significant methodological limitations and weaknesses that affect their validity and reliability. There is a lack of clear definitions for the medications included in combination therapy and those used as the reference category for comparison. Many studies simply refer to “combination therapy” and “other antidiabetic drugs,” leading to ambiguity. Moreover, different reference categories were used for comparison with metformin or combination therapy. For instance, one study compared metformin alone or in combination with other antidiabetic medications that did not include metformin [37], while another compared metformin alone to other antihyperglycemic agents, including metformin used in combination therapy [30]. Adding to it is the inconsistent definition of independent variables and the variation in their collection times across studies. These inconsistencies not only hinder comparisons between studies but also undermine the external validity of the findings, limiting their generalizability.

The lack of efforts to ensure the similarity of groups and to identify and address potential confounder factors brought possible bias. For example, Zhang *et al.* [14] divided their sample into individuals <65 years and ≥65 years and reported twenty-five statistically significant differences between the two groups in twenty-nine variables analysed. This aligns with the scientific literature that has shown that the prevalence of cardiovascular disease increases with age [64], CKD is more common among older individuals [65], and multimorbidity is also more prevalent in older adults, strongly associated with increased healthcare utilization and costs [66, 67]. This information, along with the quality assessment results, raises concerns about the internal validity of the studies and suggests a high risk of bias in the findings.

Despite these internal and external validity concerns, some observed clinical practices deviate from established guidelines and scientific literature. This misalignment highlights an urgent need to bridge the gap between clinical practice and evidence-based recommendations. Furthermore, more robust studies are essential, particularly those that emphasize external validation and minimize the risk of bias. These studies should facilitate direct comparisons across research, providing a more reliable basis for developing evidence-based recommendations grounded in high-quality data.

## Strengths and limitations

As far as we know, this is the first systematic review to map all factors driving physicians to choose metformin or combination therapy as first-line treatment. This review offers an exhaustive overview of the scientific literature, as no time or language restrictions were imposed, and grey literature was also included. Furthermore, compared to Mahmoud *et al.* [68], who conducted a meta-analysis of factors influencing antidiabetic drug prescribing for T2DM, including initiation therapy, this review expands the scope by incorporating nineteen additional studies.

All studies assessing the outcomes of interest were included, regardless of the reference group used for comparison. This approach allowed for a broad range of comparisons but also introduced heterogeneity into the analysis. Moreover, the weak evidence in the main findings due to the low quality of the studies, which integrated this systematic review, should not be overlooked.

## Conclusion

This systematic review identified several factors associated with metformin and combination therapy as first-line therapies in T2DM, revealing clinical practices not aligned with evidence-based medicine. However, few studies have focused on combination therapy, assessing physician-related factors, and even fewer have compared metformin with newer drugs, such as SGLT2i. Additionally, the studies included in the review exhibited low certainty evidence. Therefore, a robust methodology is needed to bring scientific evidence. As a result, further research is required to address these weaknesses and potential biases, provide stronger evidence, and ultimately support the findings reported in this review.

## Data Availability

All relevant data are within the manuscript and its Supporting Information files.

## Acknowledgements

We would like to thank Mandy Ryan and Michael Abbott, who supervised MH at the Health Economics Research Unit at the University of Aberdeen, for their help in developing this study, which is integrated into her master’s thesis.

## Supporting information

**S1 Table**: Search query on the PubMed (Medline)

**S2 Table**: Search query on the Scopus.

**S3 Table**: Search on the Web of Science.

## Notes

### Competing Interest Statement

The authors have declared no competing interest.

### Funding Statement

This study did not receive any funding

