## Supplementary material for "Factors influencing the prescription of first-line treatment for type 2 diabetes mellitus: a systematic review": S1 Table

S1 Table: Search query on the PubMed (Medline)

| | |
|---|---|
| #1 | (type 2 diabetes mellitus [Mesh Terms]) OR ("type II diabet*") OR (non-insulin-dependent) OR ("type 2 diabet*") |
| #2 | (time-to-treatment [Mesh Terms]) OR (initial[Title/Abstract]) OR (initiation*[Title/Abstract]) OR ("newly diagnosed"[Title/Abstract]) OR ("new diagnosis"[Title/Abstract]) OR (newly therapy[Title/Abstract]) OR (drug-naïve[Title/Abstract]) OR ("new initiator*"[Title/Abstract]) OR ("first line"[Title/Abstract]) OR (first-line [Title/Abstract]) OR ("first prescription") OR ("first therapy") OR ("treatment initiat*") OR ("initial therapy") OR ("new user*") |
| #3 | #1 and #2 |
| #4 | (hypoglycemic agents [Mesh Terms]) OR ("glucose lowering" [Title/Abstract]) OR (glucose-lowering [Title/Abstract]) OR (antihyperglycaemic [Title/Abstract]) OR (antihyperglycemic [Title/Abstract]) OR (OADs) OR (metformin) OR (sulfonylurea*) OR (alpha-glucosidase inhibitor*) OR (thiazolidinedione*) OR (dipeptidyl peptidase-4 inhibitor*) OR (DPP-4) OR (sodium-glucose cotransporter 2 inhibitor*) OR (SGLT2) NOT (insulin [Title]) |
| #5 | (choice behavior [Mesh Terms]) OR (practice patterns, physicians [Mesh Terms]) OR (drug prescription [Mesh Terms]) OR ("factors associat*" [Title/Abstract]) OR (predict* [Title/Abstract]) OR (physician* choose [Title/Abstract]) OR (pattern* [Title/Abstract]) OR ("prescribing pattern*") OR (patient-related) OR (physician-related) |
| #6 | ("factor* influencing" [Title/Abstract]) OR ("prescribing preference*" [Title/Abstract]) OR ("prescriber factor*" [Title/Abstract]) OR ("prescription factor*" [Title/Abstract]) OR (characteristic* patient*[Title/Abstract]) OR ("clinical factor*" [Title/Abstract]) OR (determinate [Title/Abstract]) OR (determinant* [Title/Abstract]) OR (influence [Title/Abstract]) OR (influencing [Title/Abstract]) OR (indicate [Title/Abstract]) OR (indicator* [Title/Abstract]) |
| #7 | #5 OR #6 |
| #8 | #3 AND #4 AND #7 |
