## Supplementary material for "Factors influencing the prescription of first-line treatment for type 2 diabetes mellitus: a systematic review": S2 Table

S2 Table: Search query on the Scopus.

| |
|---|
| (INDEXTERMS("type 2 diabetes mellitus") OR TITLE-ABS("type 2 diabet*") OR TITLE-ABS("type II diabet*") OR TITLE-ABS("non-insulin-dependent"))<br>AND<br>(INDEXTERMS(time-to-treatment) OR TITLE-ABS(initial) OR TITLE-ABS(initiation*) OR TITLE-ABS("newly diagnosed") OR TITLE-ABS("newly therapy") OR TITLE-ABS(drug-naïve) OR TITLE-ABS("first line") OR TITLE-ABS("first-line") OR TITLE-ABS (initiat*) OR TITLE-ABS("first prescription") OR TITLE-ABS("treatment initiat*") OR TITLE-ABS("initial choice") OR TITLE-ABS("initial therapy") OR TITLE-ABS(new user*) OR TITLE-ABS("new initiat*"))<br>AND<br>(INDEXTERMS(hypoglycemic agents) OR TITLE-ABS("glucose lowering") OR TITLE-ABS(glucose-lowering) OR INDEXTERMS("oral antidiabetic drugs") OR TITLE-ABS(antihyperglyc?emic) OR TITLE-ABS(metformin) OR TITLE-ABS(sulfonylurea*) OR TITLE-ABS("alpha-glucosidase inhibitor*") OR TITLE-ABS(thiazolidinedione*) OR TITLE-ABS("dipeptidyl peptidase-4 inhibitor*") OR TITLE-ABS(DPP-4) OR TITLE-ABS("sodium-glucose cotransporter 2 inhibitor*") OR TITLE-ABS(SGLT2) AND NOT TITLE(insulin))<br>AND<br>(INDEXTERMS("choice behavior") OR INDEXTERMS("drug prescription") OR TITLE-ABS("factors associat*") OR TITLE-ABS(predict*) OR TITLE-ABS(predictor*) OR INDEXTERMS("medication preferences") OR TITLE-ABS("factor* influencing") OR TITLE-ABS("prescribing preference*") OR TITLE-ABS("prescriber factor*") OR TITLE-ABS("prescription factor*") OR TITLE-ABS("prescribing criteria") OR TITLE-ABS("patient* characteristic*") OR TITLE-ABS(characteristic* PRE/1 patient*) OR TITLE-ABS("clinical factor*") OR TITLE-ABS("clinical indicator*") OR TITLE-ABS(determinate) OR TITLE-ABS(determinant*) OR TITLE-ABS(influence) OR TITLE-ABS(influencing) OR TITLE-ABS(indicate) OR TITLE-ABS(indicator*)) |
