## Supplementary material for "Factors influencing the prescription of first-line treatment for type 2 diabetes mellitus: a systematic review": S3 Table

S3 Table: Search on the Web of Science.

| |
|---|
| (TS=((“type 2 diabetes mellitus”) OR (“type 2 diabet*”) OR (“type II diabet*”) OR (non-insulin-dependent))) AND (TS= ((“initial combination therapy”) OR (“initial therapy”) OR (“newly diagnosed”) OR (drug-naïve) OR (“first line”) OR (first-line) OR (initiat*) OR (“first prescription”) OR (“treatment initiation”) OR (“new user*”) OR (newly therapy))) AND ((TS=((antidiabetics) OR (“glucose lowering”) OR (glucose-lowering) OR (“oral antidiabetic drugs”) OR (“oral antidiabetes drugs”) OR (antihyperglyc?emic)) OR ALL=((metformin) OR (sulfonylurea*) OR (alpha-glucosidase inhibitor*) OR (thiazolidinedione*) OR (“dipeptidyl peptidase-4 inhibitor*”) OR (DPP-4) OR (“sodium-glucose cotransporter 2 inhibitor*”) OR (SGLT2)) NOT TI=(insulin))) AND (TS=((“choice behavior”) OR (“practice patterns”) OR (“drug prescription”) OR (“factors associat*”) OR (predict*) OR (predictor*) OR (“treatment choices”) OR (“medication preferences”) OR (“factor* influencing”) OR (“prescribing preference*”) OR (“prescribing practice*”) OR (“prescribing criteria”) OR (“prescriber factor*”) OR (“prescription factor*”) OR (“patient characteristic”) OR (characteristic* patient*) OR (“clinical factor*”) OR (“clinical indicator*”) OR (determinate) OR (determinant*) OR (influence) OR (influencing) OR (indicate) OR (indicator*))) |
